# Drug-coated versus conventional balloons to improve recanalization of a coronary chronic total occlusion after failed attempt

**DOI:** 10.1101/2023.07.10.23292478

**Authors:** Ignacio J. Amat-Santos, Giorgio Marengo, Luiz F. Ybarra, Jose Antonio Fernández-Diaz, Ander Regueiro, Alejandro Gutiérrez, Javier Martín-Moreiras, Juan Pablo Sánchez-Luna, Jose Carlos González-Gutiérrez, Clara Fernandez-Cordon, Manuel Carrasco-Moraleja, Stéphane Rinfret

## Abstract

**Background:** Chronic total occlusion (CTO) plaque modification (CTO-PM) is often used for unsuccessful CTO interventions.

**Methods:** Multicenter, prospective study including consecutive patients with failed CTO recanalization. At the end of the failed procedure, patients received either conventional (CB) or drug-coated balloon (DCB) or at the operator’s discretion for CTO-PM and underwent new attempt of CTO recanalization ∼3 months later.

**Results:** A total of 55 patients were enrolled (DCB: 22; CB 33), with a median age of 66 years. Median J-score was 3 and CCS angina class III-IV was present in 40% of the patients. After the first CTO-PCI attempt no in hospital cardiac deaths were registered, with 3.6% rates of in-hospital myocardial infarction. The success rate of the second CTP PCI attempt was 86.8%, with periprocedural complication rate of 5.7% and without difference between DCB and CB groups. Compared with CB, in the DCB group, the second CTO-PCI required a shorter median fluoroscopy time (33 vs 60min, p<0.001), lower contrast volume (170 vs 321cc, p<0.001) and lower radiation dose (1.7 vs 3.3Gy, p<0.001). At 1-year follow up outcomes were comparable between the 2 strategies, target lesion failure occurred in 5.7% and major adverse cardiovascular events in 11.2%.

**Conclusions:** PM after CTO recanalization failure is safe and warrants high success rates when 2^nd^ attempt is performed. A DCB strategy for CTO-PM does not seem to ensure higher success or better clinical outcomes, but its use was associated with simpler staged procedures.

## INTRODUCTION

Chronic coronary total occlusions (CTOs) are highly prevalent during coronary angiography (∼10% of the patients) with almost one third of them presenting indication for percutaneous treatment (1,2). However, these procedures can be very challenging and present a success rate below 80% globally (3), as opposed to 95% in non-CTO percutaneous coronary interventions (PCI). Failed recanalization is associated not only to worse quality of life but also to a reduced survival in the follow up; for this reason, constant technical efforts are required to simplify the revascularization and increase the success rate (4–8). Highly experienced centers might present better results but frequently face more challenging cases and perform repeated procedures in up to 20% of the patients (3).

CTO plaque modification technique (CTO-PM) (9) is nowadays used as the last resource during CTO failed recanalization or when the remaining strategies are considered too high risk to modify the morphology of the CTO plaque, restore antegrade flow, and facilitate subsequent lesion recanalization (10). The technique usually consists of an intentional dilation of CTO plaque (either intraluminal or subintimal), from its proximal cap to the distal one, with a conventional semi- or noncompliant angioplasty balloon (CB), also called “STAR without stenting”. Final control angiography typically shows multiple dissection planes connecting the proximal and distal true lumen.

The use of drug-coated balloons (DCB) has largely demonstrated a high success rate in positively impacting vessel remodeling and thus reducing the atherosclerotic plaque burden (11–13). In particular the use of paclitaxel has been related with a significant regression of atherosclerotic plaque, leading to a significant increase in post-PCI lumen diameter (13). For this reason, some authors have suggested that CTO-PM with DCB may facilitate a staged procedure by promoting better vessel healing (14). The aim of this multicenter registry is to provide an updated overview of CTO-PM use, and to assess if CTO-PM with DCB might improve procedural and clinical outcomes compared with CB.

## METHODS

### Design and study population

This is a multicenter study with prospective recruitment (online database). Between October/2018 and October/2021, all CTO recanalization procedures that were failed and uncomplicated in a first attempt in six institutions were included in the study after providing informed consent. All institutions had a well-settled CTO program with >100 CTO cases per year. Allocation to receive paclitaxel-coated balloon versus CB was based on the operators’ discretion. All patients were followed up to 12-month after repeated PCI. The study was sponsored by Fundación Epic in form of an education non-conditioned grant and approved by all local ethics committees. Patients provided signed informed consent.

### CTO plaque modification technique or *investment* technique

Once the index procedure is considered unsuccessful for any reason, an angioplasty balloon was advanced intra- or extra-plaque as distal as possible into the occlusion, ideally involving the distal cap, and was inflated at nominal pressure for 1 minute. A 1:1 balloon-vessel size ratio was recommended. The choice between a conventional balloon (semi- or non-compliant) and a CDB was made according to operators’ discretion. A paclitaxel-coated Pantera Lux® balloon (Biotronik AG, Switzerland) was used in all cases allocated to the DCB group. At the end of the index procedure, use of intravascular ultrasound (IVUS) was encouraged by the investigators. Stenting was not performed during CTO-PM.

### Study endpoints

Primary endpoint was to describe procedural and clinical outcomes of CTO-PM and to assess the impact of this technique on the second index procedure performed ∼3 months after the initial one. Procedural duration, complexity (radiation, contrast administration) and complication rate during second index procedure were reported. In-hospital adverse events were showed, along with outcomes at 1-year follow up, including death, stroke, myocardial infarction (MI), target lesion revascularization, contrast-induced nephropathy (CIN) and changes in the quality of life reflected by Canadian Cardiovascular Society (CCS) angina class.

Secondary endpoints were to compare procedural success rates of the second index procedure according to the type of balloon used for the initial CTO-PM (DCB vs CB).

### Statistical analysis

Categorical variables are presented as frequencies and comparisons between groups were performed using the chi-square or the Fisher exact test. Continuous variables are expressed as median [25th-75th interquartile range] and analyzed using Mann-Whitney U test. Normal distribution was checked with Shapiro-Wilks test and q-q plots. Differences in baseline parameters across centers were evaluated and excluded. Kaplan Meier survival analysis were performed and compared with log-Rank test to determinate impact of DCB and CB in the mortality and the combined end-point of adverse events. All tests were 2 sided at the 0.05 significance level. Statistical analysis was conducted with R 3.6.3 and figures were produced using the package ggplot.

## RESULTS

### Study population

A total of 531 CTOs were treated within the study duration. Of them, 39 (7.3%) were failed with no re-attempt and in 55 patients (10.3%) – our study population – a failed CTO PCI was followed by investment technique (PM) and re-attempt. PM was performed with paclitaxel-DCB in 22 patients (40%) and with CB in 33 patients (60%). Main baseline characteristics are described in Table 1. Median age was 66 [58.5-69] years, 80% were males, and J-CTO score was ≥ 2 in 84.1% of the patients. CCS angina class III-IV was present in 40% (Figure 1) of the patients and main comorbidities included diabetes mellitus in 27.3% or chronic kidney disease in 16.4%. Median LVEF was 60 [50-60]%, 78.2% had a prior PCI and 5.5% prior coronary bypass graft surgery. Regarding coronary disease distribution, left main was involved in 7.3% and the target CTO lesion involved more often the RCA (50.9%), followed by the LAD (41.8%). There were no significant differences in baseline clinical and angiographic characteristics between CB and DCB groups.

**Figure 1.**
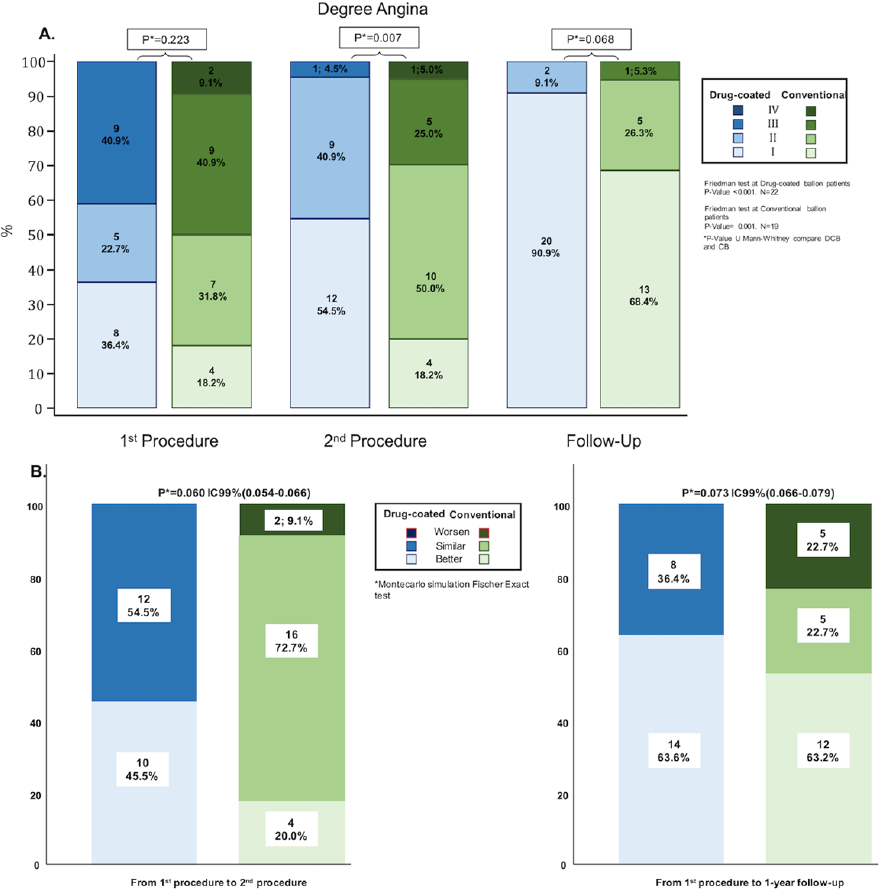
Changes in Canadian Cardiovascular Society angina class from baseline to follow up according to the use of conventional or drug-coated balloon for Chronic Total Occlusion plaque modification technique. A: Degree of angina at each time-point. B: Changes in the degree of angina.

**Table 1.**
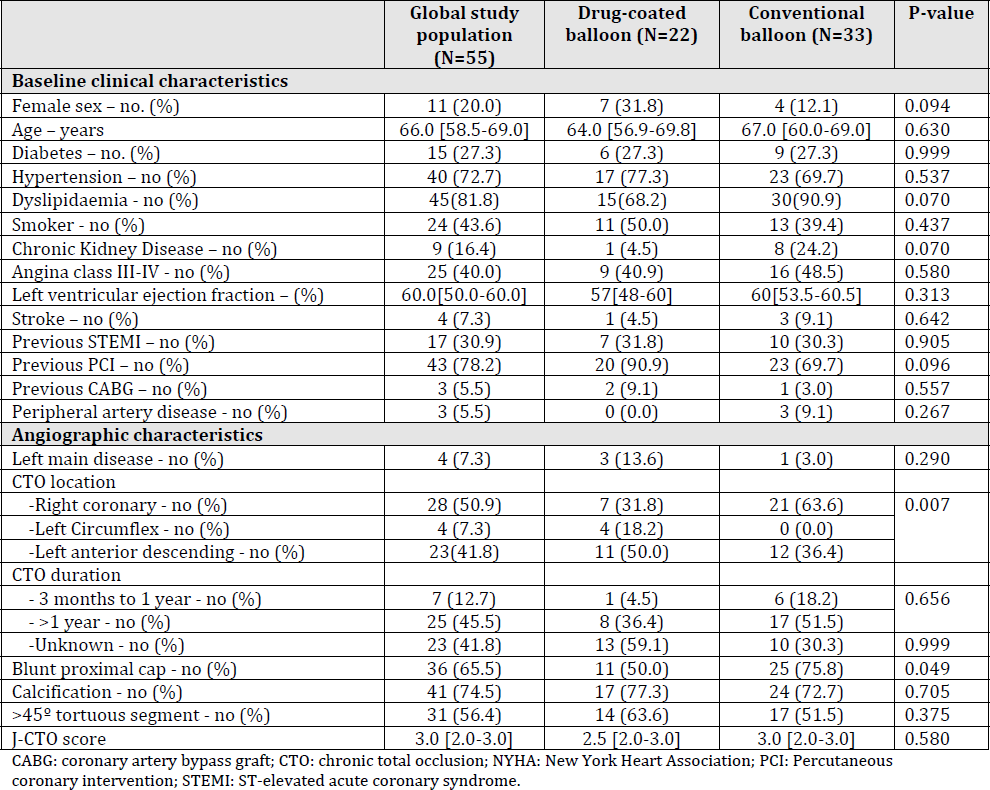
Baseline and during index procedure characteristics of the global study population and according to type of angioplasty group.

### Clinical and procedural outcomes between first and second index procedures

During the first recanalization attempt of the CTO, radial/bi-radial access was used in 23.6% of the patients, femoral/bi-femoral in 29.1% and hybrid (femoral/radial) in 47.3%. Antegrade techniques (54.5%) were the most common approach. Median fluoroscopy time was 57 min [IQR: 39-72] with a median total radiation dose of 4.3 Gy [IQR: 2.6-6.0] and contrast volume of 262 ml [IQR: 210-410].

After the first failed CTO procedure, 6 patients (10.9%) experienced in-hospital complications. Among them no in-hospital deaths were recorded, and myocardial infarction occurred in 3.6% of the patients. Pericardial effusion and acute kidney injury were uncommon (1.8% and 3.6%, respectively). Outcomes between first and second CTO recanalization attempt are reported in Table 2. Cardiac death occurred in 3.6% of the cases (2 patients) and in 5.7% a myocardial infarction was recorded. No significant differences between CB and DCB groups were noticed between the first and the second CTO-PCI attempts in terms of MACE, but there was a trend to greater symptoms relief in patients treated with DCB (Figure 1).

**Table 2.** Index procedure results, in-hospital and inter-procedural outcomes.

|  | **Global study population (N=55)** | **Drug-coated balloon (N=22)** | **Conventional balloon (N=33)** | **P-value** |
| --- | --- | --- | --- | --- |
| **Procedural aspects (Index procedure)** |  |  |  |  |
| Success rate - no % |  |  |  | - |
| Approach |  |  |  |  |
| -antegrade - no % | 30(54.5) | 12 (54.5) | 18 (54.5) | 0.225 |
| -retrograde - no % | 5(9.1) | 1 (4.5) | 4 (12.1) |  |
| -hybrid – no (%) | 20(36.4) | 9 (40.9) | 11 (33.3) |  |
| Access |  |  |  |  |
| -Radial/Bi-radial – no (%) | 13(23.6) | 5 (22.7) | 8 (24.2) | 0.260 |
| -Femoral/Bi-femoral – no (%) | 16(29.1) | 9 (40.9) | 7 (21.2) |  |
| -Radial/Femoral – no (%) | 26(47.3) | 8 (36.4) | 18 (54.5) |  |
| Fluoroscopy time (min) | 57.0 [39.0-72.0] | 47.0 [32.0-71.0] | 61.0 [42.5-72.0] | 0.406 |
| Contrast volume (cc) | 262 [210-410] | 253 [198-400] | 272 [216-430] | 0.673 |
| Total radiation (Gy) | 4.3[2.6-6.0] | 4.8[2.9-6.1] | 3.2[2.4-5.5] | 0.198 |
| IVUS performed during first attempt – no (%) | 22(40.0) | 8 (36.4) | 14 (42.4) | 0.653 |
| **In-hospital outcomes (Index procedure)** |  |  |  |  |
| In-hospital complications – no (%) | 6(10.9) | 4 (18.2) | 2 (6.1) | 0.204 |
| In-hospital death – no (%) | 0 (0.0) | 0 (0.0) | 0 (0.0) | 0.999 |
| Myocardial infarction – no (%) | 2(3.6) | 1 (4.5) | 1 (3.0) | 0.999 |
| CABG – no (%) | 0 (0.0) | 0 (0.0) | 0 (0.0) | 0.999 |
| Stroke – no (%) | 0 (0.0) | 0 (0.0) | 0 (0.0) | 0.999 |
| Pericardial effusion – no (%) | 1(1.8) | 1 (4.5) | 0 (0.0) | 0.400 |
| Acute renal failure – no (%) | 2(3.6) | 0 (0.0) | 2 (6.1) | 0.511 |
| Other in-hospital complications – no (%) | 4(7.3) | 4 (18.2) | 0 (0.0) | **0.021** |
| **Clinical events between Index and 2nd Index Procedure** |  |  |  |  |
| Death – no (%) | 2(3.6) | 0 (0.0) | 2 (6.1)* | 0.511 |
| Cardiac death – no (%) | 2(3.6) | 0 (0.0) | 2 (6.1)* | 0.511 |
| Myocardial infarction – no (%) | 3(5.7) | 0 (0.0) | 3 (9.7) | 0.258 |
| CABG – no (%) | 0 (0.0) | 0 (0.0) | 0 (0.0) | 0.999 |
| Stroke – no (%) | 0 (0.0) | 0 (0.0) | 0 (0.0) | 0.999 |
| New revascularization – no (%) | 2(6.3) | 1 (8.3) | 1 (5.0) | 0.999 |

\*Two patients in the conventional balloon group died before repeated procedure, one due to ventricular arrhythmia and other due to intracranial bleeding following fortuitous fall.
CABG: coronary artery bypass graft; IVUS: Intravascular ultrasonography.

### Peri-procedural and in-hospital outcomes after second index procedure

Median time between first and second recanalization attempt was 77 [70-86] days with no differences between groups. Success rate in the second attempt was 86.8% and a hybrid approach (with both antegrade and retrograde techniques) was performed in 52.9% of the cases. A radial/femoral access was chosen in 56.6% of the patients, and a biradial access in 17.2%. Median fluoroscopy time was 47 min [IQR: 31-62], total radiation dose was 2.6 Gy [IQR: 1.7-3.6] and contrast volume was 241 ml [IQR: 152-338]. Periprocedural complications included 5.7% myocardial infarctions, 13.7% cases complicating with acute kidney injury, and no procedural mortality. The in-hospital outcomes also excluded further mortality and are summarized in Table 3.

**Table 3.** Second index procedure results, in-hospital and long-term outcomes.

|  | **Global study population (N=55)** | **Drug-coated balloon (N=22)** | **Conventional balloon (N=33)** | **P-value** |
| --- | --- | --- | --- | --- |
| **Procedural aspects (second attempt)** |  |  |  |  |
| Procedural success – no (%) | 46(86.8) | 18 (81.8) | 28 (90.3) | 0.431 |
| Approach |  |  |  |  |
| -antegrade - no % | 15(29.4) | 4 (18.2) | 11 (37.9) | 0.175 |
| -retrograde - no % | 9(17.6) | 6 (27.3) | 3 (10.3) |  |
| -hybrid – no (%) | 27(52.9) | 12 (54.5) | 15 (51.7) |  |
| Access |  |  |  |  |
| -Radial/Bi-radial – no (%) | 9(17.2) | 3(13.6) | 6(19.4) | 0.929 |
| -Femoral/Bi-femoral – no (%) | 14(26.4) | 6(27.3) | 8(25.8) |  |
| -Radial/Femoral – no (%) | 30(56.6) | 13(59.1) | 17(54.8) |  |
| Fluoroscopy time – minutes | 47[31-62] | 33[23-44] | 60[49-69] | **<0.001** |
| Contrast volume – ml | 241[152-338] | 170[119-219] | 321[231-401] | **<0.001** |
| Total radiation (Gy) | 2.6[1.7-3.6] | 1.7[1.2-2.1] | 3.3[2.6-4.1] | **<0.001** |
| Plaque modification – no (%) | 5(9.6) | 3 (13.6) | 2 (6.7) | 0.639 |
| Complications – no (%) | 3(5.7) | 0 (0.0) | 3 (9.7) | 0.258 |
| Periprocedural death – no (%) | 0 (0.0) | 0 (0.0) | 0 (0.0) | 0.999 |
| Cardiac tamponade – no (%) | 2(3.8) | 0 (0.0) | 2 (6.5) | 0.505 |
| Stroke – no (%) | 0 (0.0) | 0 (0.0) | 0 (0.0) | 0.999 |
| **In-hospital outcomes (second attempt)** |  |  |  |  |
| Complications – no (%) | 7(13.2) | 2 (9.1) | 5 (16.1) | 0.686 |
| Death – no (%) | 0 (0.0) | 0 (0.0) | 0 (0.0) | 0.999 |
| Myocardial infarction – no (%) | 3(5.7) | 1 (4.5) | 2 (10.0) | 0.999 |
| CABG – no (%) | 0 (0.0) | 0 (0.0) | 0 (0.0) | 0.999 |
| Stroke – no (%) | 0 (0.0) | 0 (0.0) | 0 (0.0) | 0.999 |
| Pericardial effusion – no (%) | 3(5.7) | 0 (0.0) | 3 (9.7) | 0.258 |
| Acute renal failure – no (%) | 7(13.7) | 2 (9.1) | 5 (17.2) | 0.684 |
| Other in-hospital complications – no (%) | 0 (0.0) | 0 (0.0) | 0 (0.0) | 0.999 |
| **One-year outcomes** |  |  |  |  |
| Death at 12 months – no (%) | 3(5.7) | 0(0.0) | 3(9.1) | 0.267 |
| Myocardial infarction – no (%) | 4(7.3) | 1 (4.5) | 3 (9.1) | 0.999 |
| CABG – no (%) | 0(0) | 0 (0.0) | 0 (0.0) | 0.999 |
| Stroke – no (%) | 0(0) | 0 (0.0) | 0 (0.0) | 0.999 |
| Target lesion revascularization – no (%) | 3(5.7) | 2 (9.1) | 1 (3.0) | 0.557 |
| MACE^ at 12 months – no (%) | 6(11.4) | 1 (4.5) | 5 (15.2) | 0.384 |
| MACE^ + Acute renal failure – no (%) | 14(27.4) | 3 (13.6) | 11 (33.3) | 0.100 |

\*Two patients in the conventional balloon group died before repeated procedure.
CABG: coronary artery bypass graft; ^MACE: Major adverse cardiovascular events (All-cause mortality, myocardial infarction, stroke, target lesion revascularization).

Compared with CB group, second index procedure was shorter in the DCB-group, requiring shorter fluoroscopy time (33 [23-44] vs 60 [49-69] min, p<0.001), lower contrast volume (170 [119-219] vs 321 [231-401] cc, p<0.001) and lower radiation dose (1.7 [1.2-2.1] vs 3.3 [2.6-34.1] Gy, p<0.001) as summarized in Figure 2. Other in-hospital outcomes were comparable between the 2 groups.

**Figure 2.**
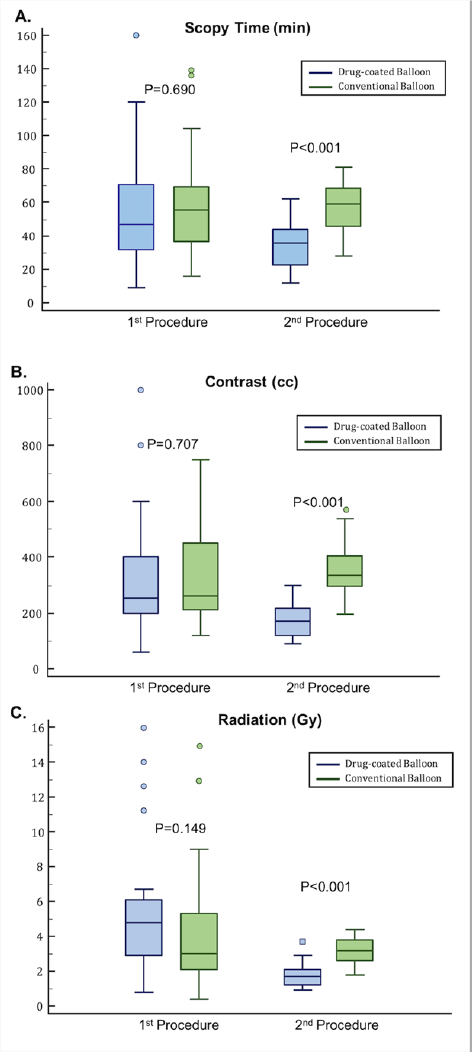
Fluoroscopic time, contrast volume and radiation used during first and second index procedures according to the use of conventional or drug-coated balloon.

### One-year outcomes after second index procedure

At 1-year follow up MACE rate was 11.4% of the patients, with 5.7% cardiovascular deaths, 7.3% myocardial infarctions, and 5.7% target lesion revascularization rate. No significant differences in 1-year outcomes were observed between CB and DCB groups (Figure 3). The combined endpoint of MACE and acute kidney injurty was numerically higer in the CB group (33.3% vs 13.6%, p=0.1), not reaching the statistical significance though.

**Figure 3.**
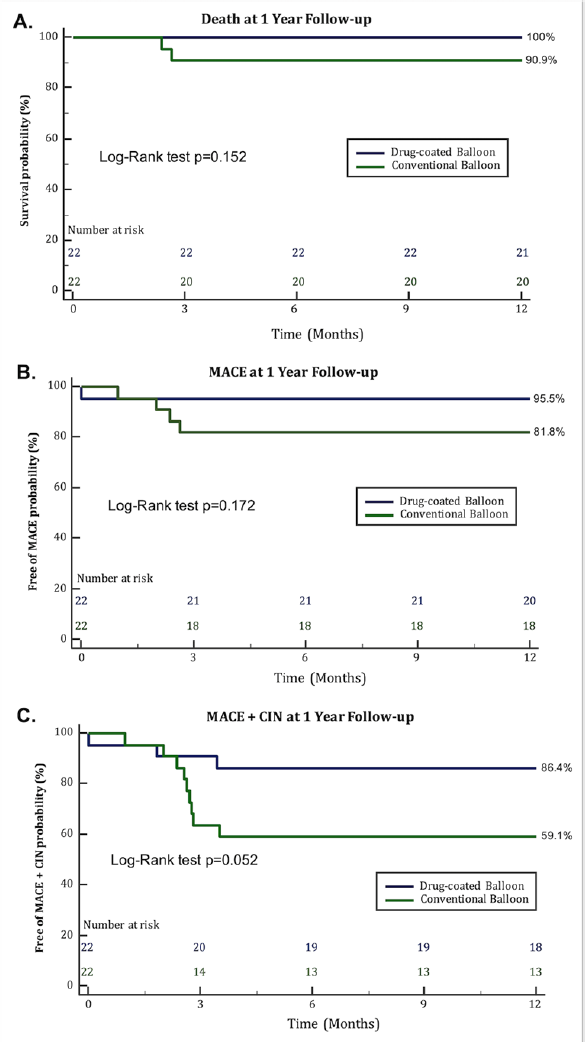
One-Year mortality (A) and combined end-points (B: Death + Stroke + Myocardial infarction + Target lesion revascularization); C: Same as B + Contrast-induced nephropathy) according to the use of conventional or drug-coated balloon.

## DISCUSSION

In this multicenter registry we provide an updated overview of clinical and procedural outcomes of CTO-PM technique. Additionally, we aimed to investigate whether a DCB strategy (with paclitaxel coated balloon) in the first failed CTO-PCI attempt might lead to better success rates of the second attempt than CB-based strategy. The main findings of this analysis are as follows: 1) CTO-PM technique is a safe approach and ensures low periprocedural complication and high success rates in the second attempt (∼87%). 2) A DCB strategy, compared with a CB one, does not provide better clinical outcomes in terms of MACE, but is related with greater improvement in anginal status, suggesting that the DCB might promote a better vessel healing. 3) Although the success rate of the second attempt was comparable irrespective of the balloon used, the use of DCB was associated to shorter and simpler procedures, with lower contrast and radiation doses. 4) There were no cases of eccentric remodeling or extensive dissections in the second index procedure suggesting lack of local paclitaxel toxicity when delivered in the intra- or extra-plaque space.

### CTO plaque modification technique for CTO recanalization

Initially described by Vo MN et al (15), CTO-PM technique for increasing success recanalization rate, also known as *investment* technique or subintimal plaque modification, has been receiving growing importance and is currently included in the global CTO crossing algorithm as a standard technique if other approaches fail (16). According to the data from the PROGRESS-CTO registry (10), around 20% of the cases in highly experienced centers had unsuccessful recanalization attempts and, of them, CTO-PM was performed in only 13%. To remark, in these cases success rate of subsequent CTO PCI was 83% and was particularly high in those cases performed ≥60 days after the index procedure (94%). Complication rates related to this technique have been constantly reported as low, in agreement with our findings. As previously reported by Hirai T et al (17), apart from facilitating ulterior recanalization attempts, CTO-PM might also relief angina symptoms by itself, and this occurred also in our patients. Interestingly, in our analysis the symptoms relief was even greater when a DCB *investment* strategy was planned, suggesting that even if new attempts are considered unlikely, CTO-PM with DCB might be taken into consideration as a “palliative” strategy.

### Local impact of paclitaxel delivery in the extra-plaque space

Even if drug-eluting stents are still considered the therapy of choice for interventional treatment of coronary artery disease, in the last years drug coated balloons (DCB) have been more and more proposed as valid alternatives (18, 19). Nowadays, DCBs are a widely accepted therapy for coronary in-stent restenosis (18) and for small vessel coronary lesions (19), but promising results are ongoing in other scenarios and especially for the treatment of coronary lesions without prior stent implantation.

Because of its unique properties, Paclitaxel is largely used for DCB coating. This drug is typically used for the treatment of different types of cancer due to its anti-proliferative effects, based on directly affecting cellular mitosis by increasing tubulin polymerization (20). However, regarding its effect on coronary disease and despite its largely use for coronary intervention devices, its mechanisms of regression of atherosclerosis are complex and not fully understood. Through these unraveled mechanisms, also in the complex scenario of CTO the use of Paclitaxel facilitated the revascularization in a subsequent procedure after a failed first attempt likely by modifying plaque morphology and size beyond a merely mechanical effect of balloon dilation (13).

It is known that vessels treated with DCBs usually show good plaque stabilization at follow up along with larger lumen gain that those obtained by CB suggesting a positive remodeling on the local atherosclerotic plaque (21). Such effect might be also obtained if the drug is delivered in the extra-plaque space, leading to the persistence and the expansion of the healing connections achieved by the CTO-PM, communicating the proximal and distal true lumen either subintimally or intraplaque. Interestingly in our study, the procedure length of the second attempt was significantly shorter when a DCB strategy was firstly chosen despite the lack of anatomical differences suggesting that in the context of CTO-PCI, intraplaque or subintimal DCB dilation can improve device deliverability and distal wiring in a second attempt simplifying second CTO-PCI.

### Long-term effect of CTO-PM with DCB

Despite the safety demonstrated by paclitaxel-coated balloons for the treatment of coronary disease (18), some concerns were raised regarding its use for the treatment of peripheral arterial disease (19) due to an increase in global mortality after the first year of follow-up according to Katsanos K et al (19). It has been also previously suggested that DCB with paclitaxel might predispose to the appearance of arterial aneurisms (19). In our small casuistic, we did not observe any. A longer follow-up with invasive or non-invasive coronary angiography might be needed to clarify this aspect but the lack of adverse vessel remodeling in our research suggest lack of negative structural of systemic effects. In fact, Scheller B et al have previously demonstrated a reduction in 3-year follow up all-cause mortality when DCBs are used for the treatment of coronary disease but its use is still not extended due to acute angiographic results with early recoil and concerns about dissections causing acute vascular occlusions (18). These aspects are not applicable for its use during CTO-PM technique, but still some concerns regarding paclitaxel effect might exist. The low solubility of paclitaxel prevents immediate dissolution following contact with blood, limits loss of paclitaxel from balloon, modulates efficacy and toxicity by limiting the maximum achievable concentration, and contributes to the long-lasting efficacy. Paclitaxel concentrations in the vessel wall immediately after DCB inflation are far above its solubility representing the sum of dissolved (i.e., the pharmacologically active drug) plus solid crystalline paclitaxel, which serves as a reservoir but does not exhibit toxicity or pharmacological effects (18). The dose administered by a coronary DCB is approximately 750 times lower compared with systemic cancer therapy, challenging the plausibility of a drug effect when local therapies do not have systemic effects.

The main limitation of this study is its small sample size that might limit the power to detect differences between CDB and CB groups regarding periprocedural success rate and complications. However, this is one of the largest prospective registries available focused on the use of CTO-PM following CTO recanalization failure and with systematic second attempt. Although baseline characteristics between DCB and CB groups were very similar, the lack of randomization is a limitation and the eventual beneficial effect of DCBs in simplifying staged procedures must be interpreted as hypothesis generating. Only one specific brand of paclitaxel DCB was used in our analysis, but multiple conventional balloons were used, therefore external validity of our finding will require further investigation. Cost-effectiveness studies might be also required in this setting.

## CONCLUSIONS

CTO-PM technique after a failed CTO recanalization is safe and warrants a high success rate when a second attempt is performed. In this context, the use of DCBs compared with CBs did not increase procedural success, but was associated to greater early clinical improvement, and provided shorter staged procedures duration, and lower contrast and radiation doses. Further studies with greater sample size and a randomized design are needed to confirm the beneficial effect of DCB for CTO-PM.

## FUNDING STATEMENT

The study was sponsored by Fundación Epic in form of an institutional educational grant (Epic-BTK20).

## Data Availability

Data are available for other investigators under request.

## ABBREVIATIONS

CTO: Chronic coronary total occlusion.
CTO-PM: Chronic coronary total occlusion plaque modification.
CB: Conventional balloon.
DCB: Drug-coated balloon.
IVUS: Intravascular ultrasound.
LAD: Left anterior descending artery.
LCx: Left circumflex artery.
MI: Myocardial infarction.
PCI: Percutaneous coronary intervention
RCA: Right coronary artery
TIMI: Thrombolysis in Myocardial Infarction.

## AKNOWLEDGMENTS

None

## CONFLICTS OF INTEREST

Drs. Amat-Santos and Rinfret are proctors for Boston Scientific. There are no other conflicts of interest to declare by any author.

## Notes

### Clinical Trial

NCT05158686

### Funding Statement

The study was sponsored by FundaciÃ^3^n Epic in form of an institutional educational grant (Epic-BTK20).

### Author Declarations

CEIm from Valladolid Este provided approval and was transferred to the ECs from the other participating institutions.

